# Development of a behavioral model for the use of psychosocial support services by cancer survivors: A grounded theory pilot study

**DOI:** 10.1101/2025.07.31.25332471

**Authors:** Manae Hirayama, Daisuke Kasai, Ken-ichi Tabei

## Abstract

This pilot study explored the factors influencing the theoretical use of psychosocial support services among cancer survivors not currently using those services. It used the Fogg Behavior Model as a theoretical framework. A web-based questionnaire survey was conducted over one month using snowball sampling through social networking services of patient organizations, targeting cancer survivors aged 40 years and older. Free-text responses about support needs from participants who were not using psychosocial support were analyzed using a grounded theory approach. The texts were coded based on the three Fogg Behavior Model elements: “triggers” to initiate behavior, “motivation” to engage in behavior, and “ability” that hinders behavior. The structure serving as the starting point for behavior was modeled. The results revealed that many factors were based on emotional values, such as expectations and hopes for support. “Motivation” indicated the need to build trust with supporters and to organize feelings. “Ability” identified that barriers included self-sufficiency and difficulties verbalizing feelings. “Trigger” required supporters to show human qualities, and for there to be clear pathways to consultation and creative publicity methods. These findings suggest that acquiring psychological values such as reassurance, trust, and hope strongly motivates the use of psychosocial support. Designing appropriate support pathways based on cancer survivors’ behaviors and developing effective cognitive strategies are important ways to promote the future use of psychosocial support services.

## Introduction

In recent years, advances in cancer drugs and treatments have improved the survival rates for many types of cancer, leading to an increase in the number of cancer survivors [1]. It is estimated that there are more than 50 million cancer survivors worldwide [2], including approximately 3.41 million in Japan [3]. Cancer survivors, regardless of whether they are still receiving treatment, may experience feelings of anxiety or depression, known as emotional distress. Approximately 30% of individuals continue to experience these symptoms even after treatment ends [4].

Emotional distress affects mental health and also has physical and behavioral impacts, leading to an overall decrease in the quality of life [5]. Psychosocial support is considered effective in improving quality of life [6], and providing this support is crucial [7]. In Japan, efforts are being made to promote the use of cancer consultation support centers by providing accurate information [8], but awareness is only 55.1%, and usage remains at 21.1%, indicating that these services are not being fully used [9]. Additionally, patients who need psychosocial support but do not receive it experience greater distress than those who do [10]. According to a survey by an insurance company, about 60% of cancer survivors stated they “hesitate to ask for help” [11].

Other studies have identified stigma attached to psychosocial support, such as the desire to keep personal affairs to themselves, or not wanting to have personal problems managed by others, as well as concerns about whether support is available to meet individual needs [12][13][14].

Another study investigating barriers to availability of psychosocial care services highlighted reasons such as healthcare providers prioritizing treatment or medical procedures over psychosocial care, and a shortage of professionals to provide this support. Even in high-income countries, the psychosocial needs of cancer survivors are generally not fully met [15]. Previous studies have reported low utilization rates, user trends, and the characteristics of those who do not need psychosocial support. However, it is not clear which specific behavioral factors would improve utilization rates. This study aimed to identify the factors that would encourage cancer survivors who do not currently use psychosocial support services to start to do so. We used a descriptive approach, with the Fogg Behavior Model (FBM) [16] as a theoretical framework to explain the use of psychosocial support services. The FBM is a theoretical model that indicates the three necessary elements to trigger behavior: motivation, ability, and trigger [16]. Using this framework clarifies the specific behavioral factors related to the use of psychosocial support by cancer survivors, systematizes the design and evaluation of interventions, and facilitates a common understanding among stakeholders [17]. This makes it easier to examine the effectiveness of intervention measures.

Several recent studies have applied the FBM to cancer. For example, one study used a design based on the FBM to make proposals to improve compliance with a liver cancer screening intervention using a mobile health approach, focused on high-risk groups [18]. A pilot randomized controlled trial of a mobile app promoting genetic counseling use among ovarian cancer patients showed improvements in knowledge, self-efficacy, and family communication [19]. These suggest that applying the FBM to the use of psychosocial support by cancer survivors is also likely to be effective. This study aimed to clarify the specific motivational, ability-related, and trigger factors affecting the use of psychosocial support by cancer survivors. It therefore qualitatively analyzed the factors associated with willingness to use support services using the FBM, and proposed strategies to improve utilization rates. It is expected to contribute to the development of effective intervention strategies to improve use of support services.

## Methods

### Materials

Using Google Forms and snowball sampling, the questionnaire was distributed to cancer survivors aged 40 years or older through the social networking services of multiple patient groups over a one-month period (May 1 to May 31, 2024). The questionnaire included (1) questions about the use of psychosocial support for cancer patients, with three questions for those who answered “yes” and two questions for those who answered “no,” and (2) one open-ended question about expectations of psychosocial support.

We adopted a detailed methodological approach based on the FBM and analyzed the survey results. This approach enables a comprehensive understanding of the factors influencing cancer survivors’ use of psychosocial support services. The sample size for this pilot study was determined based on the following criteria. For continuous variables [20] [21], 12 participants were required in each group. We also required a total of 36 participants to ensure a 95% confidence interval (0.85–0.95) with a 5% margin of error [22].

### Statistical analysis

The basic attributes, use of psychosocial support, and situation from diagnosis to the present were analyzed using descriptive statistics. For the content analysis of psychosocial support needs among those not using counseling support, we used the grounded theory approach [23][24] to structure the psychosocial needs of cancer patients and the actual state of support. The free descriptions were open-coded. We defined code families and concepts based on the three elements of the FBM, motivation, ability, and trigger. Theoretical coding [25] was used to model the structure underlying the behavior (Table 1).

**Table 1.**
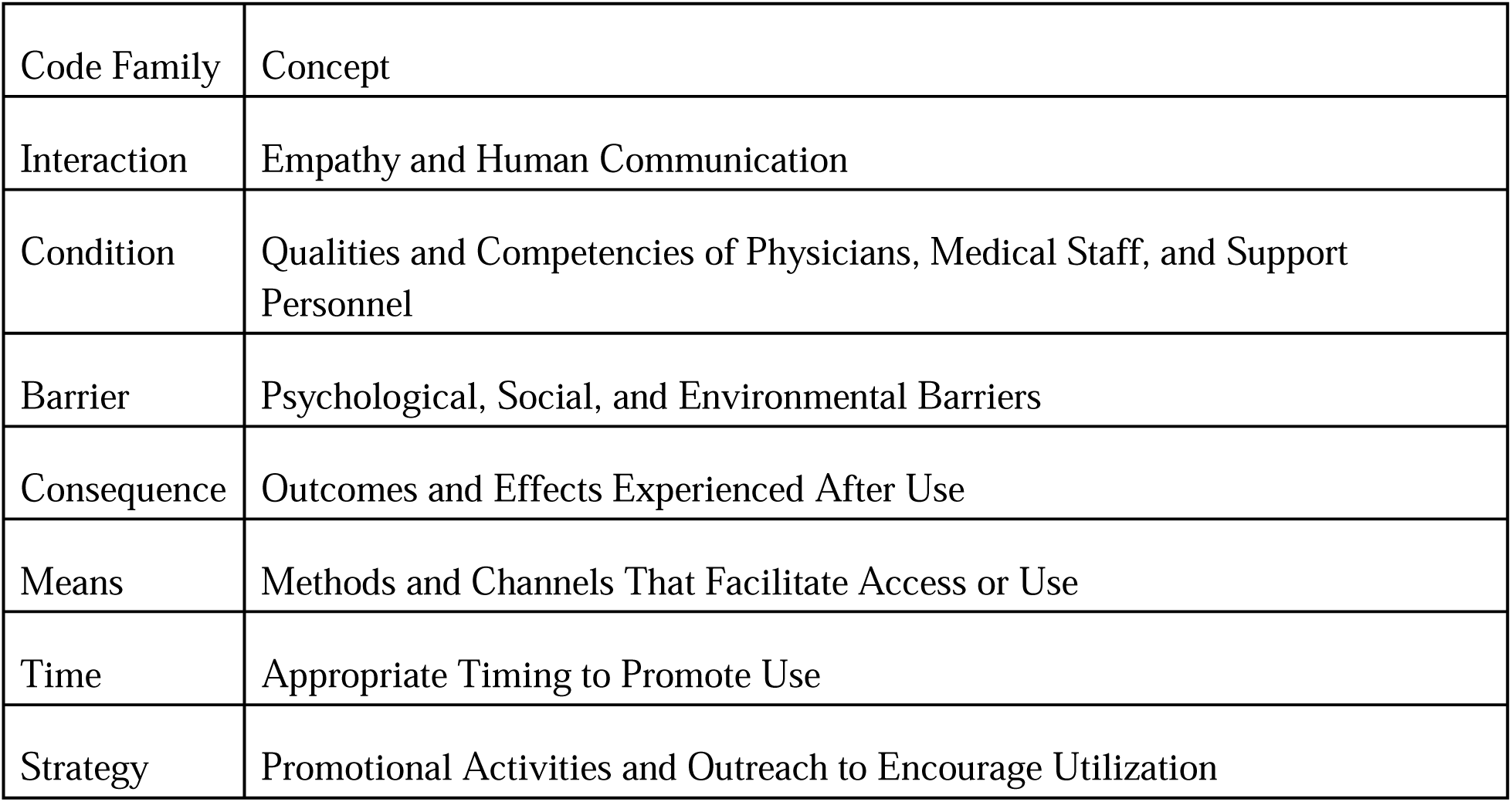
Code families used to categorize the requests of cancer survivors not using psychosocial support.

| Code Family | Concept |
| --- | --- |
| Interaction | Empathy and Human Communication |
| Condition | Qualities and Competencies of Physicians, Medical Staff, and Support Personnel |
| Barrier | Psychological, Social, and Environmental Barriers |
| Consequence | Outcomes and Effects Experienced After Use |
| Means | Methods and Channels That Facilitate Access or Use |
| Time | Appropriate Timing to Promote Use |
| Strategy | Promotional Activities and Outreach to Encourage Utilization |

The first author was responsible for the survey design and initial coding of free descriptions. During the analysis, the data were reviewed and discussed multiple times with co-authors experienced in medicine and patient research to eliminate biased perspectives and unilateral interpretations of the data. R Version 4.3.2 was used for descriptive statistics, and MAXQDA was used for the theoretical coding of descriptive data.

### Ethical approval

This study was approved by the Research Safety and Ethics Committee of the Tokyo Metropolitan University of Industrial Technology (Approval No. 23020). The questionnaire survey was conducted in accordance with the Declaration of Helsinki and relevant guidelines, and informed consent was obtained from all respondents. All response data were anonymized.

## Results

### Characteristics of participants

Responses were obtained from 63 participants. The average age was 55.5 years, with 43 women (68.3%) and 20 men (31.7%) included in the study (Table 2). Overall, 32 people (50.8% said that they had used psychosocial support services. The reasons for use were as follows: “Had questions or concerns” (15 people, 46.9%), “Wanted information about medical or welfare services” (6 people, 18.8%), “Wanted to learn about treatment or care options” (three people, 9.4%), “Needed support from family or those around them” (one person, 3.1%), and “Other” (seven people, 21.9%). The psychosocial support services used included the Cancer Consultation Support Center (18 people), cancer salons or patient groups (10 people), telephone services (two people), apps or online services (one person), and consulting medical professionals (one person).

**Table 2.**
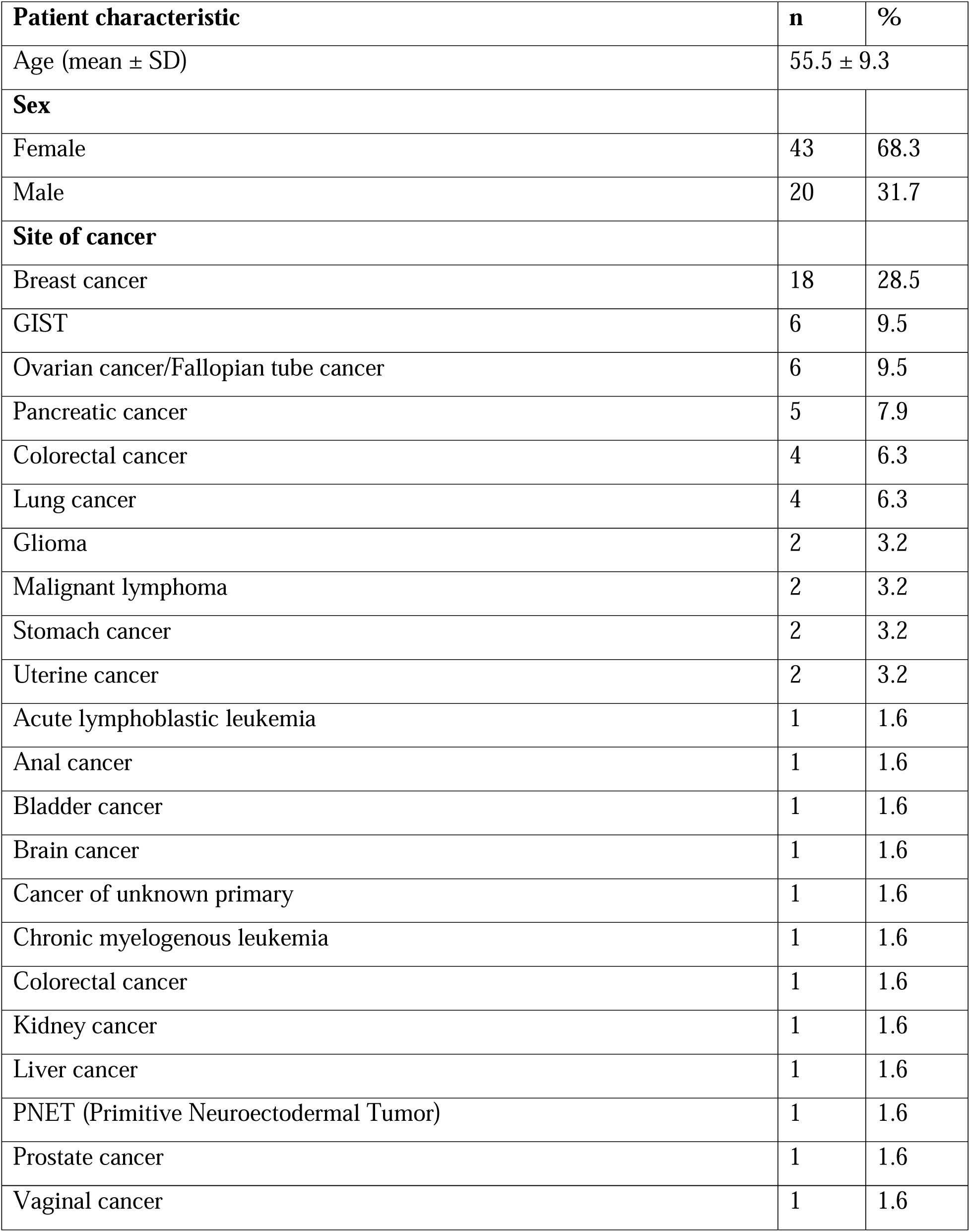
Basic attributes of participants.

In contrast, 31 (49.2%) participants said that they did not use these services. The reasons for not using them were: “I sufficiently understand my current situation” (four people, 12.9%), “I research and find information myself” (six people, 19.4%), “I do not feel the need for counselling support” (five people, 16.1%), “The support provided by medical professionals is sufficient” (four people, 12.9%), and “Other” (13 people, 41.9%) (Table 3).

**Table 3.** Trends in the use of consultation support.

|  | n | % |
| --- | --- | --- |
| Have you ever used cancer counseling support? |  |  |
| Yes | 32 | 50.8 |
| No | 31 | 49.2 |
| What motivated you to seek cancer counseling support? |  |  |
| Had questions or concerns | 15 | 46.9 |
| Wanted to know about treatment and care options | 3 | 9.4 |
| Needed support from family or others | 1 | 3.1 |
| Wanted information about medical and social services | 6 | 18.8 |
| Other | 7 | 21.9 |
| What type of cancer counseling support do you use? |  |  |
| Cancer Consultation Support Center | 1 | 1.6 |
| Cancer salons and patient groups | 18 | 28.6 |
| Telephone services | 10 | 15.9 |
| Mobile apps and online services | 2 | 3.2 |
| Other | 1 | 1.6 |
| Why did you not use counselling support? |  |  |
| The present situation is fully comprehended. | 3 | 9.7 |
| The present author is conducting independent research and gathering information. | 6 | 19.4 |
| It is not perceived to be necessary to seek counselling or support. | 5 | 16.1 |
| The level of support provided by my healthcare provider is adequate. | 4 | 12.9 |
| Other | 13 | 41.9 |

### Barriers and facilitators to the use of psychosocial support

This section focuses on insights gained from open-ended responses by individuals not using psychosocial support. We also qualitatively analyzed the specific factors that influenced the use of support. The results were categorized into themes such as “Motivation”, which underpins behavior, “Ability”, which enables or inhibits behavior; and “Trigger”, which prompts behavior. We also included “Interaction”, “Conditions”, “Outcomes”, “Means”, “Strategies”, “Barriers” and “Time” (Table 4). Of the 32 open-ended responses, 16 (50.0%) were about motivation, 13 (40.6%) about triggers, and two (6.3%) about ability. Fig 1 shows a model of psychosocial support use behavior created by adapting the FBM using these results. The model indicates that when the ability barrier influencing behavior is lower, it is more likely that a trigger will succeed in driving action. Conversely, when the barrier is higher, it is more likely that a trigger will fail to drive action. This suggests that when motivation, the driving force behind the behavior, is high, a trigger is more likely to drive action, whereas when motivation is low, it is more likely to fail to do so (Fig 1).

**Fig 1.**
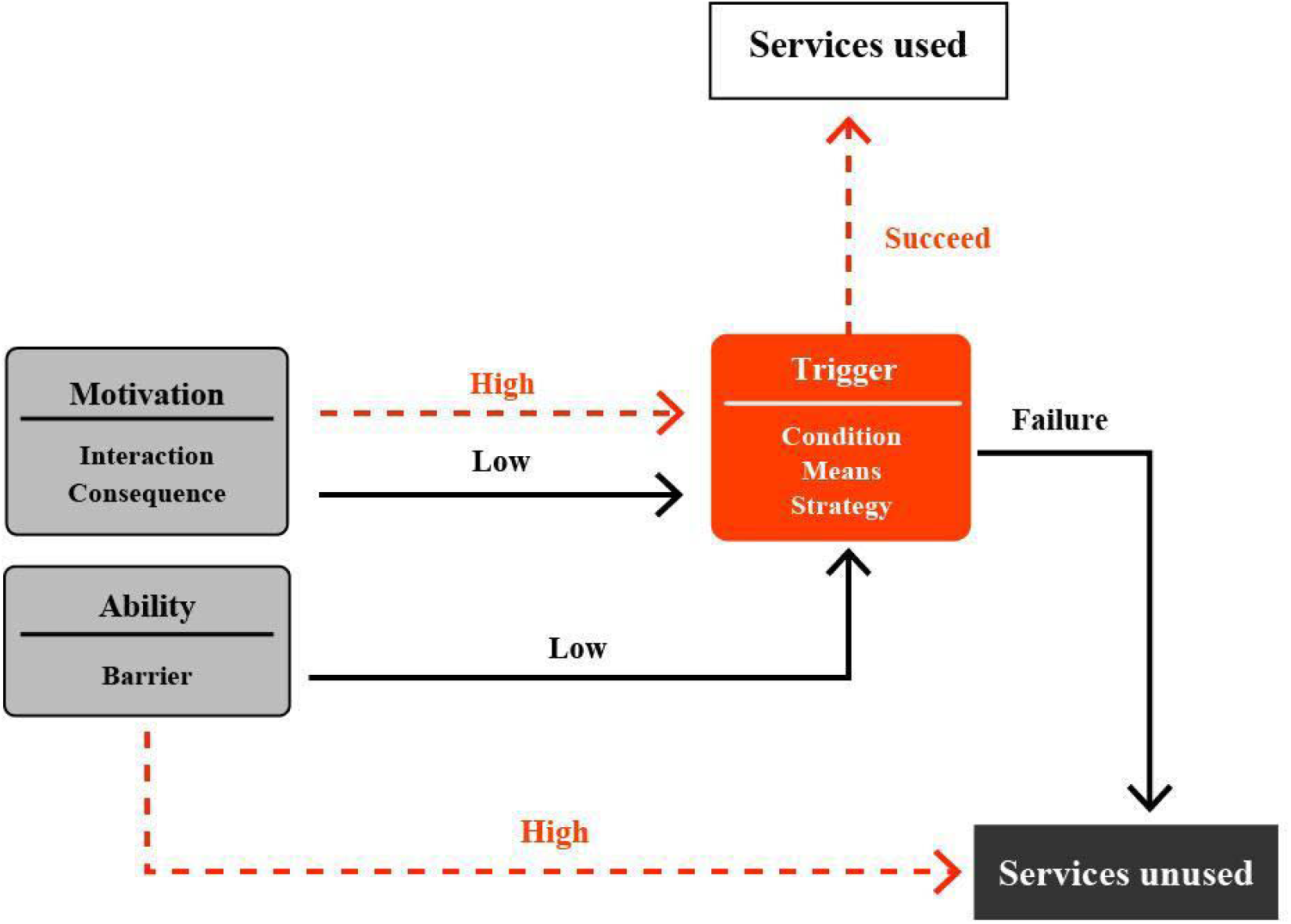
Behavioral model of psychosocial support use based on Fogg’s Behavior Model

**Table 4.** The coded text and themes.

| Sex | FBM | Theme | Text |
| --- | --- | --- | --- |
| Female | Trigger | Condition | It helps when someone with empathy reaches out to you. |
| Female | Trigger | Condition | It feels easier to use when someone initiates the conversation. |
| Female | Trigger | Means | I want support that understands my work and workplace situation. |
| Female | Trigger | Time | It's helpful if someone listens before the consultation or while waiting. |
| Female | Trigger | Means | It would be great to have online consultations via Zoom or similar platforms. |
| Female | Trigger | Means | Support that helps search for necessary information together would be beneficial. |
| Female | Trigger | Means | I wanted to connect with other patients who have similar experiences. |
| Female | Trigger | Means | An orientation session right after diagnosis would be very helpful. |
| Male | Trigger | Condition | It helps to get advice from a trustworthy expert when you're unsure. |
| Male | Trigger | Strategy | I'd appreciate having booklets and flyers readily available to take home. |
| Male | Trigger | Strategy | I'd like to see videos or interviews of other patients using consultation services. |
| Male | Trigger | Means | I think a counseling service could relieve stress by allowing people to share concerns. |
| Male | Trigger | Strategy | Promoting it in a way that makes me think, “Maybe I should try it,” would help. |
| Female | Ability | Barrier | I’ve had a vague unease I can’t put into words, so I don’t know how to seek help. |
| Male | Ability | Barrier | I tend to rely on looking things up myself and don’t seek help from others. |
| Female | Motivation | Consequence | I want to ask questions I find hard to bring up with my doctor. |
| Female | Motivation | Consequence | Routine appointments make it hard to raise issues; I want to share even minor concerns. |
| Female | Motivation | Interaction | I’ve felt lonely since diagnosis, and I hope someone can support me through it. |
| Female | Motivation | Consequence | I need help maintaining a positive mindset and emotional balance. |
| Female | Motivation | Consequence | I wanted to learn how to support my child emotionally while managing my own feelings. |
| Female | Motivation | Consequence | I’d like referrals or info on hospitals for preventing recurrence or metastasis. |
| Female | Motivation | Interaction | I appreciate when someone listens closely and helps organize my thoughts and feelings. |
| Female | Motivation | Interaction | A place where they truly listen to your story is valuable. |
| Female | Motivation | Interaction | Support should be personalized based on individuals and their family context. |
| Female | Motivation | Interaction | Support systems should vary to match the patient’s stage and specific needs. |
| Male | Motivation | Interaction | A place where hope for life can be nurtured through counseling is important. |
| Male | Motivation | Consequence | I want guidance on how to distance myself from people who aren't helpful. |
| Male | Motivation | Consequence | I'd like to know if remission is possible and how to achieve it. |
| Male | Motivation | Consequence | I want comprehensive information, including clinical trials. |
| Male | Motivation | Consequence | I lack information due to limited explanations from my doctor, so I want more details. |
| Male | Motivation | Consequence | I want a one-stop place to talk about everything—from medical info to daily concerns. |
| Female | Motivation | Interaction | Rather than qualifications, supporters should be chosen based on their actual abilities. |

The following section discusses each theme in more detail.

#### Theme 1: Motivation—security and hope brought about by interaction and outcomes

The primary motivations for using psychosocial support were centered around building trust through interaction and obtaining a sense of security and hope. For example, one user expressed expectations about the support provider’s character and ability to empathize with users:

“*We should select people with ability, rather than just those with qualifications (such as social workers or nurses) or suitability*.” (Interaction/Motivation)

Expectations for psychosocial support included the provision of information or assistance with decision-making and also organizing feelings and gaining a sense of reassurance.

“*When I want to ask something that’s difficult to bring up with the doctor, I can ask*.” (Outcome/Motivation) These points indicate a desire to make a human connection and have some hope for the future, beyond just the “information” or “skills” provided by psychosocial support.

#### Theme 2: Barriers to action (ability): obstacles to use caused by difficulty articulating feelings and a tendency toward self-reliance

Two statements corresponded to “Ability”, and they shared themes such as “difficulty putting feelings into words” and “hesitation to rely on others”.

“*I’ve constantly carried a vague sense of unease that I can’t put into words, so I don’t know how to ask for advice or seek support*.” (Female/Barrier/Ability) “*I tend to think I understand things after researching them myself, so I don’t really try to rely on others*.” (Male/Barrier/Ability)

The former expresses frustration at the “difficulty explaining in words”, and the latter reveals hesitation because of a self-reliant attitude. Neither participant rejected psychosocial support itself, but both were in a psychological state in which it is hard to find an “opportunity” to access support. This highlights the complexity of “Ability” within the FBM.

#### Theme 3: Trigger: promoting usage behavior through presentation of conditions, means, and strategies

The trigger to encourage behavior required not only “conditions” and “means”, but also “strategically designed pathways”.

##### Conditions

“*It should be someone with human depth*.” (Condition/Trigger)

“*To receive advice based on expertise from someone trustworthy*.” (Condition/Trigger) The person who responds and the attitude they take was an important starting point in making participants feel that it would be okay to consult with that person.

##### Means

“*It would be good to have a mechanism where orientation is received immediately after diagnosis*.” (Means/Trigger)“*Support where necessary information can be searched together*.” (Means/Trigger)

Means help to develop a pathway to action, and supportive guidance acts as a trigger.

##### Strategies

“*If the service was publicized, I might want to use it*.”(Strategy/Trigger)

“*I want to see videos or interviews showing what a consultation support center looks like from the patient’s perspective*.” (Strategy/Trigger)

This indicates that “ingenious measures to encourage potential users” are crucial to promote the use of psychosocial support.

## Discussion

### Principal results

In this study, we hypothesized that cancer survivors who do not use psychosocial support services experience challenges such as stigma about seeking support and a lack of trust in these services. Based on the FBM, we investigated the factors related to the use of psychosocial support. The analysis revealed that many respondents held emotional values such as expectations and hopes for psychosocial support. However, factors such as “a lack of appropriate pathways” and “an environment that does not create opportunities for action” emerged as barriers to its use in practice.

### Comparison with prior work

First and foremost, motivation focused on building trust through interactions and obtaining a sense of security and hope as a result. Women, in particular, emphasized expectations of support to enable “emotional sorting” and developing a sense of security. Previous studies have also reported that female cancer survivors tend to feel anxious [26] and have a strong tendency to seek empathy and support for their feelings [27][28], which is consistent with our results. In contrast, men emphasized the need for information about clinical trials and the remission period of their illness, focusing less on psychological support. Previous studies have also shown that men have a low demand for psychological support [29], which is consistent with our findings.

We also obtained insights about barriers to action related to ability, such as difficulties expressing emotions. This highlighted a psychological state of having no opportunity or not wanting to rely on others. The tendency for self-sufficiency may be attributed to the Dunning– Kruger effect [30]. However, those who self-rate their health literacy[31] as high [32] often do not realize the inadequacy of their understanding. This tendency has been reported in other studies [33][34], and was also reflected in our results. The inability to verbalize emotions, known as inhibited self-disclosure, has been noted as a factor hindering help-seeking behavior in patients with ovarian cancer [35], which is consistent with our findings.

Triggers included aspects such as the humanity of supporters, clarification of consultation or counselling procedures, and promotional activities to encourage awareness. These findings suggest the need to balance a safe access environment with mechanisms that promote action when introducing psychosocial support. Surveys targeting patients with advanced cancer have also reported the importance of ensuring accessibility to support [36][37], which our findings confirm. Previous research also suggests the need for flexible support tailored to conditions and intentions rather than uniform responses for all patients [38]. Our findings indicate the potential to promote use of support services more effectively by presenting a combination of elements that trigger actions. This could serve as concrete guidelines for designing behavior-changing interventions for psychosocial support.

### Strengths and limitations

This study involved several limitations. First, because it was a pilot study conducted via a web based questionnaire, selection bias may have occurred. Future research should address this issue by employing larger, more representative samples and a variety of data collection methods.

Second, although we outlined a clear framework for discussing limitations, practical implications, and future research directions, our findings may not generalize to all cancer survivors who do not use psychosocial support services.

Third, our analysis was restricted to the descriptive content provided in the one time survey responses, limiting the depth of insight. Complementary qualitative studies—such as in depth interviews or ethnographic observations—are needed to enrich these findings.

Fourth, because the Google Form link was shared on third party platforms without trackable URLs, we could not perform click through analysis or calculate a response rate. Future surveys should use trackable links to obtain precise engagement metrics.

Finally, building on these results, subsequent research should develop and evaluate practical interventions—such as tailored counseling services and informational tools—to optimize psychosocial support for cancer survivors.

## Conclusion

In this study, we used the FBM to systematically organize the factors that promote the use of psychosocial support. This has provided insights that may be useful for understanding the initial design and intervention points for behavioral change in the future. In particular, psychosocial support is not merely a place to provide information. Based on the identified importance of motivation and triggers, it will be necessary to focus on enhancing trust and hope through individualized communication strategies and clearly defined pathways for initiating support for patients.

## Data Availability

Due to the nature of this study, participants did not consent to the public sharing of data, so supporting data is not available.

## Acknowledgments

We would like to express our deep gratitude to the patient organizations and respondents who cooperated with the recruitment and survey for this study. We also thank Melissa Leffler, MBA, from Edanz (https://jp.edanz.com/ac) for editing a draft of this manuscript.

