## Supplementary material for "Development of a behavioral model for the use of psychosocial support services by cancer survivors: A grounded theory pilot study": JARS-Qual_Checklist (from another version of this preprint: version 1, 2025-07-31)

| Section | Subsection | JARS-Qual Requirement | Check section |
|---|---|---|---|
| Title Page | Title | Identify key issues/topic under consideration. | Title |
| | Author Note | • Acknowledge funding sources or contributors.<br>• Acknowledge conflicts of interest, if any | Acknowledgments/Funding Statement/Competing Interests |
| | Abstract | • State the problem/question/objectives under investigation.<br>• Indicate the study design, including types of participants or data sources, analytic strategy,main results/findings, and main implications/significance.<br>• Identify five keywords. | Abstract |
| Introduction | Research Problem | • Frame the problem or question and its context.<br>• Review, critique, and synthesize the applicable literature to identify key issues/debates/ theoretical frameworks in the relevant literature to clarify barriers, knowledge gaps, or practical needs. | Introduction |
| | Study Objectives/Aims/Research Goals | • State the purpose(s)/goal(s)/aim(s) of the study.<br>• State the target audience, if specific.<br>• Provide the rationale for fit of design used to investigate this purpose/goal (e.g., theory building, explanatory, developing understanding, social action, description, highlighting social practices).<br>• Describe the approach to inquiry, if it illuminates the objectives and research rationale (e.g., descriptive, interpretive, feminist, psychoanalytic, postpositivist, critical, postmodern,<br>constructivist, or pragmatic approaches). | Introduction |
| | Research Design Overview | • Summarize the research design, including data-collection strategies, data-analytic strategies, and, if illuminating, approaches to inquiry (e.g., descriptive, interpretive, feminist,<br>psychoanalytic, postpositivist, critical, postmodern, constructivist, or pragmatic approaches).<br>• Provide the rationale for the design selected. | Method/Materials |
| | Researcher Description | • Describe the researchers' backgrounds in approaching the study, emphasizing their prior understandings of the phenomena under study (e.g., interviewers, analysts,<br>or research team).<br>• Describe how prior understandings of the phenomena under study were managed and/or influenced the research (e.g., enhancing, limiting, or structuring data collection and analysis). | Method/Materials |

| | | |
|---|---|---|
| Participants or Other Data Sources | • Provide the numbers of participants/documents/events analyzed.<br>• Describe the demographics/cultural information, perspectives of participants, or characteristics of data sources that might influence the data collected.<br>• Describe existing data sources, if relevant (e.g., newspapers, internet, archive).<br>• Provide data repository information for openly shared data, if applicable.<br>• Describe archival searches or process of locating data for analyses, if applicable. | Method/Materials |
| Researcher–Participant Relationship | • Describe the relationships and interactions between researchers and participants relevant to the research process and any impact on the research process (e.g., was there a relationship prior to research, are there any ethical considerations relevant to prior relationships). | Method/Materials |
| Recruitment Process | Describe the recruitment process (e.g., face-to-face, telephone, mail, email) and any recruitment protocols.<br>• Describe any incentives or compensation, and provide assurance of relevant ethical processes of data collection and consent process as relevant (may include institutional review board approval, particular adaptations for vulnerable populations, safety monitoring).<br>• Describe the process by which the number of participants was determined in relation to the<br>study design.<br>• Provide any changes in numbers through attrition and final number of participants/sources<br>(if relevant, refusal rates or reasons for dropout).<br>• Describe the rationale for decision to halt data collection (e.g., saturation).<br>• Convey the study purpose as portrayed to participants, if different from the purpose stated.<br>The order of the recruitment process and the selection process and their contents may be determined in relation to the authors' methodological approach. Some authors will determine a selection process and then develop a recruitment method based on those criteria. Other authors will develop a recruitment process and then select participants | Method/Materials |

| | | | |
|---|---|---|---|
| Method | Participant Selection | • Describe the participants/data source selection process (e.g., purposive sampling methods,<br>such as maximum variation; convenience sampling methods, such as snowball selection; theoretical sampling; diversity sampling) and inclusion/exclusion criteria.<br>• Provide the general context for the study (when data were collected, sites of data collection).<br>• If your participant selection is from an archived data set, describe the recruitment and selection process from that data set as well as any decisions in selecting sets of participants from that data set. | Method/Materials |
| | Data Collection Procedures | • State the form of data collected (e.g., interviews, questionnaires, media, observation).<br>• Describe the origins or evolution of the data-collection protocol.<br>• Describe any alterations of data-collection strategy in response to the evolving findings or the study rationale.<br>• Describe the data-selection or data-collection process (e.g., were others present when data<br>were collected, number of times data were collected, duration of collection, context).<br>• Convey the extensiveness of engagement (e.g., depth of engagement, time intensiveness of data collection).<br>• For interview and written studies, indicate the mean and range of the time duration in the data-collection process (e.g., interviews were held for 75 to 110 min, with an average interview time of 90 min).<br>• Describe the management or use of reflexivity in the data-collection process, as it illuminates the study.<br>• Describe questions asked in data collection: content of central questions, form of questions<br>(e.g., open vs. closed). | Method/Statistical analysis |
| | Recording & Data Transformation | • Identify data audio/visual recording methods, field notes, or transcription processes used. | Method/Statistical analysis |

| | | | |
|---|---|---|---|
| | Data Analysis | • Describe the methods and procedures used and for what purpose/goal.<br>• Explicate in detail the process of analysis, including some discussion of the procedures (e.g., coding, thematic analysis) following a principle of transparency.<br>• Describe coders or analysts and their training, if not already described in the researcher description section (e.g., coder selection, collaboration groups).<br>• Identify whether coding categories emerged from the analyses or were developed a priori.<br>• Identify units of analysis (e.g., entire transcript, unit, text) and how units were formed, if applicable.<br>• Describe the process of arriving at an analytic scheme, if applicable (e.g., if one was developed before or during the analysis or was emergent throughout).<br>• Provide illustrations and descriptions of the analytic scheme development, if relevant.<br>• Indicate software, if used. | Method/Statistical analysis |

| | | | |
|---|---|---|---|
| | Methodological Integrity | • Demonstrate that the claims made from the analysis are warranted and have produced findings with methodological integrity. The procedures that support methodological integrity (i.e., fidelity and utility) typically are described across the relevant sections of a paper, but they could be addressed in a separate section when elaboration or emphasis would be helpful. Issues of methodological integrity include the following:<br>– Assess the adequacy of the data in terms of its ability to capture forms of diversity most relevant to the question, research goals, and inquiry approach.<br>– Describe how the researchers' perspectives were managed in both the data collection and analysis (e.g., to limit their effect on the data collection, to structure the analysis).<br>– Demonstrate that findings are grounded in the evidence (e.g., using quotes, excerpts, or descriptions of researchers' engagement in data collection).<br>– Demonstrate that the contributions are insightful and meaningful (e.g., in relation to the current literature and the study goal).<br>– Provide relevant contextual information for findings (e.g., setting of study, information about participants, interview question asked is presented before excerpt as needed).<br>– Present findings in a coherent manner that makes sense of contradictions or disconfirming evidence in the data (e.g., reconcile discrepancies, describe why a conflict might exist in the findings).<br>• Demonstrate consistency with regard to the analytic processes (e.g., analysts may use demonstrations of analyses to support consistency, describe their development of a stable perspective, interrater reliability, consensus) or describe responses to inconsistencies, as relevant (e.g., coders switching midway through analysis, an interruption in the analytic process). If alterations in methodological integrity were made for ethical reasons, explicate<br>those reasons and the adjustments made.<br>• Describe how support for claims was supplemented by any checks added to the qualitative<br>analysis. Examples of supplemental checks that can strengthen the research may include | Method/Statistical analysis |
